# Examining changes in sleep duration associated with the onset of the COVID-19 pandemic: Who is sleeping and who is not?

**DOI:** 10.1101/2021.04.06.21254996

**Authors:** Salma Batool-Anwar, Rebecca Robbins, Shahmir H. Ali, Ariadna Capasso, Joshua Foreman, Abbey M. Jones, Yesim Tozan, Ralph J. DiClemente, Stuart F. Quan

**Author notes:** Correspondent: Stuart F. Quan, M.D., Division of Sleep and Circadian Disorders, Brigham and Women’s Hospital, 221 Longwood Ave., Boston, MA 02115. Attestation: All authors have seen and approved this manuscript. Conflicts of Interest: Dr. Robbins has received personal fees from Denihan Hospitality, Rituals Cosmetics, byNacht, and SleepCycle. Dr. Quan serves as a consultant to Jazz Pharmaceuticals, Best Doctors and Whispersom. Drs. Batool-Anwar, Foreman, Tozan, DiClemente, and Mr. Ali, and Mmes. Capasso and Jones have no conflicts of interest.

## Abstract

**Introduction:** The COVID-19 pandemic has resulted in social isolation and reports of insomnia. However, reports of changes in sleep duration and associated factors are few.

**Methods:** Data were from an online survey of adults recruited via social media that included a question asking whether the respondent slept less or more after the onset of the pandemic. Analyses determined the association between changes in sleep duration and self reported sociodemographic and occupational information; beliefs about COVID-19; changes in sleep patterns; and responses pertaining to loneliness, anxiety, and depression.

**Results:** There were 5,175 respondents; 53.9% had a change in sleep duration. 17.1% slept less and 36.7% slept more. Sleeping more was related to greater education, being single/divorced/separated, unemployed or a student. Being retired, divorced/separated or a homemaker, and living in the Mountain or Central time zones were associated with less sleep. Beliefs that COVID-19 would result in personal adverse consequences was associated with both more and less sleep. However, the strongest associations with both more and less sleep were seen with depression, anxiety, and loneliness with adjusted odds ratios ranging from 1.92 (*95% CI* 1.67-2.21) for sleeping more and loneliness to 5.29 (*95% CI* 4.1-6.7) for sleeping less and anxiety.

**Conclusions:** Changes in sleep duration since the start of the COVID-19 pandemic were highly prevalent among social media users and were associated with several sociodemographic factors and beliefs that COVID-19 would have adverse personal impacts. However, the strongest associations occurred with worse mental health suggesting that improvements may occur with better sleep.

## Introduction

It is widely recognized that sleep is a vital psychophysiologic process for mental health. In addition to increased prevalence of mental health conditions among people with sleep disorders,^1^ a bidirectional relationship also exists between sleep and psychiatric disorders.^2^ This association is further strengthened during times of crisis from natural disasters and pandemics. Increased rates of insomnia among adolescents (38.3%) were seen after the Wenchuan earthquake of 2008^3^, and these sleep disturbances were associated with depression and post traumatic stress disorder.^4^ Similar experiences of a negative effect on mental health were seen during previous pandemics.^5^. After the MERS pandemic of 2015, high rates (47.2%) of anxiety symptoms were seen as many as six months after the isolation ended.^6^. Individuals infected with SARS and MERS also reported greater anxiety, depression, insomnia, and post-traumatic stress disorder (PTSD) during and in the post-illness stage.^7^

These experiences from past epidemics and natural disasters have raised concerns for imminent psychosocial impact following the current COVID-19 pandemic. Although SARS-CoV-2 mainly affects the respiratory system, recent reports have demonstrated neurologic symptoms related to COVID-19.^8^ There has been increased focus on neuropsychiatric effects of the virus and its relationship with impaired sleep.^7,9^.

The current pandemic has caused increased mortality, economic instability, and social isolation. The devastation of this magnitude is generating considerable burden in terms of population-wide mental health.^5,10^ Besides the direct effects of SARS-CoV-2 on the brain, the detrimental effects of lockdown and quarantine on psychosocial health also have been reported globally.^11-13^ Increased anxiety and stress have been reported among people during isolation and quarantine.^5^ Increased cases of stigmatization and discrimination during these periods of social isolation have been shown to further contribute to psychosocial stress.^14^ Additionally, the effects of home confinement on light exposure and more access to use of digital tools has been shown to have a negative impact on sleep quality.^15^

Worsening psychosocial as well as sleep health can be seen among vulnerable populations. Healthcare workers including physicians, nurses, laboratory workers, and technicians delivering care at the front line are considered the most vulnerable during a pandemic situation. A recent meta-analysis demonstrated increased rates of anxiety, depression, PTSD, and insomnia among healthcare workers affecting their well-being.^16^ They are put in an unprecedented situation by having to make difficult decisions and work under extreme pressure, both resulting in moral injury.^17^

Although several studies have reported increased depression, irritability, PTSD, and insomnia among COVID-19 survivors and healthcare workers, the data on the impact of COVID-19 on changes in sleep duration among the general population are limited. The objective of this study was to extend the results of previous studies by examining the effect of the COVID-19 pandemic on self-reported changes in sleep duration and the sociodemographic and mental health factors associated with these changes among the general population of the United States (US).

## Methods

### Participants

Recruitment for this survey has been described extensively elsewhere.^18,19^ Briefly, during April 16-21, 2020, a social-media advertisement campaign was launched on Facebook and affiliated platforms (i.e., Instagram, Facebook Messenger) to attract participation in a survey of COVID-19 knowledge, beliefs and behaviors. Previous research has shown social media to be a valid method of recruitment for health surveys, including those focused on COVID-19.^13,20^ The campaign was targeted to persons who were at least 18 years of age and residing in the United States. Interested social media users were directed to an anonymous web-based survey administered by Qualtrics (Provo, UT). Eligibility was confirmed by brief screening questions. Participants were not compensated for survey completion. All study material and procedures underwent Human Subjects Research review by the New York University Institutional Review Board and were determined to be exempt, and the need for explicit written or oral consent was also waived (IRB-FY2020–4285).

### Questionnaire

Survey development has been outlined previously.^18,19^ In summary, the survey was based on principles expressed in the Health Belief Model,^21^ a paradigm used in surveys related to other viral outbreaks.^22-24^ In addition to demographic information, the survey included a variety of questions related to the participant’s behaviors, attitudes and beliefs towards COVID-19 as well as their current mental health. Items were adapted from surveys utilized during previous infectious disease outbreaks so that they were pertinent to the COVID-19 pandemic.^25^ The survey also contained a single question regarding sleep: [“Since hearing about the Coronavirus outbreak, how have the below behaviors changed for you? Sleeping”]. Responses were elicited using a 5-point Likert scale: “Much less,” “Little less,” “Not changed,” “Little more,” and “Much more” with an additional Not Applicable response. Participants who selected Not Applicable were removed from the analysis. These responses were aggregated into 3 categories “sleeping less than usual” (combination of “Much less” and “Little less sleep”), “sleeping more than usual” (combination of “Little more” and “Much more”) and “no change in sleep.” Besides sociodemographic characteristics of the cohort, we chose additional survey items to be included in these analyses based on our *a priori* assessment that their content would be relevant to changes in sleep behavior.

Beliefs about COVID-19 assessed included perceived risk [“What do you think is your risk of getting infected with Coronavirus?”] and perceived severity [“If you were infected with Coronavirus, how severe do you think it would be?”], which were measured on a 10-point scale from “not at all likely” to “extremely high risk” and “extremely severe,” respectively. Responses to perceived risk and severity were re-coded so responses of “not at all” to the scale midpoint (5) represented “low” beliefs about risk and severity, while responses above the scale mid-point (6) to 10 (Very Severe) were re-coded to represent “high” beliefs about risk and severity. Responses to financial ability to quarantine [“I can financially afford to self-quarantine”], beliefs about discrimination [“Since the Coronavirus outbreak, I feel discriminated against”], beliefs that COVID-19 is deadlier than influenza [“Coronavirus is more deadly than the seasonal flu”], beliefs COVID-19 is not as big a problem as the media suggest [“Coronavirus is not as big of a problem as the media suggests”], and beliefs that COVID-19 is a bigger problem than the government suggests [“Coronavirus is a bigger problem than the government suggests”] were captured on scales from 0 “strongly disagree” to 3 “strongly agree.” Responses were dichotomized so that “strongly disagree” and “disagree” were re-coded to indicate “low” beliefs or concerns, while “disagree” or “strongly disagree” were re-coded to represent “high” beliefs or concerns, with the exception of financial ability to quarantine, which was re-coded so that “strongly agree” and “agree” were represented low financial worries and “strongly disagree” and “disagree” represented high financial worries.

Mental health was assessed using the Patient Health Questionnaire-4 (PHQ-4),^26^ an ultra-brief screening scale for depression and anxiety, which was included in the survey and modified to refer to feelings about the coronavirus outbreak over the past 7 days. Anxiety was considered present if there was a score of 3 or greater on the first 2 items of the PHQ-4. Similarly, depression was determined to be present if there was a score of 3 or greater on the last 2 items of the PHQ-4. In addition to anxiety and depression, the construct of “loneliness” was assessed using responses to the following 3 questions: “In the last 3 months, how often have your felt: 1) left out; 2) isolated; 3) that you lacked companionship.” Possible responses for all 3 questions were the following: 0) Never; 1) Often; 2) Rarely; 3) Sometimes. A cumulative score of 6 or higher was considered as indicative of “loneliness.”^27^ Additional details regarding the wording of the survey items and possible responses are provided in the supplement.

### Statistical Analysis

We estimated differences in sociodemographic characteristics, COVID-19 beliefs, and mental health by reported change in sleep (i.e., no change, versus either sleeping more than usual or less than usual). We utilized Pearson 𝒳^2^ to determine if associations existed between sociodemographic factors and the reported change in sleep. Next, to examine sleep changes by sociodemographic factors, we conducted multivariable binary logistic regressions to characterize the sociodemographic profile of those who reported sleeping more than usual and less than usual. In both cases, models used reports of “no change in sleep” as the reference group. To examine COVID-19 beliefs by sleep changes, we ran adjusted multivariable binary logistic regressions to examine reports of sleeping more and sleeping less and their relationship to COVID-19 beliefs, after adjusting for confounders (i.e., age, sex, race, presence of a child at home, urban residence, marital status, education, employment, and time zone). Finally, to examine mental health by sleep changes, we conducted adjusted binary logistic regression to examine reports of sleeping more and sleeping less and their relationship to loneliness, anxiety, and depression, controlling for the aforementioned confounders. The level of statistical significance for all models was set at 0.05. Analyses were conducted in SPSS (Version 21; Armonk, NY).

## Results

There were 5,175 participants who responded to the survey question pertaining to change in sleep. Their demographic information disaggregated by whether they reported a change in sleep or not, is shown in Table 1. Irrespective of whether they reported a change in their sleep duration, most respondents were between the ages of 50 and 69 years (26-34%), predominantly of white race (92.7%-93.5%), and more likely to be female (54.6-62.2%). A majority also reported being married (68.7%-74.1%), residing in an urban area (51.6%-55%) in the Eastern time zone (54%-57.8%), not having children in the household (71.9%-76.4%), having gone to college (88.6%-91.7%), and being employed (53.7%-58.9%).

**Table 1.** Demographic characteristics of the sample by report of any change in sleep (i.e., either reports of sleeping more or sleeping less than usual versus no change in sleep, N=5, 175)

|  |  | <u>No Change in Sleep</u> |  | <u>Any Change (Slept More or Slept Less)</u> |  | **Chi-Square** | **P-Value** |
| --- | --- | --- | --- | --- | --- | --- | --- |
| **Variables** |  | **N** | **%** | **N** | **%** |  |  |
| Age | >70 | 313 | 13.1% | 201 | 7.2% | 120.2 | 0.000 |
|  | 60-69 | 811 | 34.0% | 736 | 26.4% |  |  |
|  | 50-59 | 624 | 26.1% | 825 | 29.6% |  |  |
|  | 40-49 | 335 | 14.0% | 487 | 17.5% |  |  |
|  | 30-39 | 227 | 9.5% | 376 | 13.5% |  |  |
|  | 18-29 | 78 | 3.3% | 162 | 5.8% |  |  |
| Sex | Male | 1,078 | 45.4% | 1,048 | 37.8% | 30.8 | 0.000 |
|  | Female | 1,294 | 54.6% | 1,725 | 62.2% |  |  |
| Race | White | 2,214 | 92.7% | 2,606 | 93.5% | 9.8 | 0.007 |
|  | Black, Hispanic, Asian | 73 | 3.1% | 104 | 3.7% |  |  |
|  | Other | 101 | 4.2% | 77 | 2.8% |  |  |
| Child | No children at home | 1,825 | 76.4% | 2,004 | 71.9% | 13.5 | 0.000 |
|  | Child at home | 563 | 20.3% | 783 | 28.1% |  |  |
| Urban/Rural | Rural | 786 | 34.6% | 772 | 28.8% | 19.9 | 0.000 |
|  | Urban | 1173 | 51.6% | 1473 | 55.0% |  |  |
|  | Suburban | 315 | 13.9% | 433 | 16.2% |  |  |
| Marital Status | Married/cohabiting | 1,686 | 74.1% | 1,839 | 68.7% | 27.3 | 0.000 |
|  | Single | 316 | 13.9% | 480 | 17.9% |  |  |
|  | Widowed | 106 | 4.7% | 102 | 3.8% |  |  |
|  | Divorced/separated | 166 | 7.3% | 257 | 9.6% |  |  |
| Education | High School Diploma or GED | 257 | 11.4% | 218 | 8.3% | 14.1 | 0.000 |
|  | Some College or Higher | 1,989 | 88.6% | 2,424 | 91.7% |  |  |
| Work | Employed | 1,222 | 53.7% | 1,577 | 58.9% | 121.6 | 0.000 |
|  | Unemployed | 123 | 5.4% | 270 | 10.1% |  |  |
|  | Student | 14 | 0.6% | 47 | 1.8% |  |  |
|  | Retired | 726 | 31.9% | 547 | 20.4% |  |  |
|  | Unable to work | 92 | 4.0% | 142 | 5.3% |  |  |
|  | Housework | 97 | 4.3% | 95 | 3.5% |  |  |
| Time Zone | Pacific | 251 | 11.1% | 327 | 12.3% | 14.2 | 0.002 |
|  | Mountain | 185 | 8.2% | 178 | 6.7% |  |  |
|  | Central | 606 | 26.8% | 620 | 23.3% |  |  |
|  | Eastern | 1221 | 54.0% | 1539 | 57.8% |  |  |

Overall, 2,787 (53.9%) participants experienced a change in their sleep duration as a consequence of the COVID-19 pandemic. As shown in Table 1, younger persons were more impacted by the pandemic than older individuals. In addition, more women (62.2%) than men (37.8%) altered their sleep duration. Other demographic factors associated with a large (>3% absolute difference) change in sleep duration were living in an urban area (55.0% [changed] vs. 51.6% [unchanged]), having a child at home (28.1% [changed] vs. 20.3% [unchanged], being single (17.9% [changed] vs. 13.9% [unchanged]), or being employed (58.9% [changed] vs 53.7% [unchanged]. Conversely, living in a rural area (28.8% [changed] vs. 34.6% [unchanged]) and being retired (20.4% [changed] vs. 31.9% [unchanged]) appeared to have a large protective effect on change in sleep duration.

Associations of sociodemographic factors with sleeping less since hearing about the COVID-19 outbreak are displayed in Table 2. There were 884 (17.1%) participants who slept less than usual. Significant associated factors were younger age (<60 years), female sex, urban and suburban residential area and being divorced or separated.

**Table 2.** Binary Logistic Regression Models Examining Sociodemographic Predictors of Sleeping Less than Usual and Sleeping More Than Usual (N=5, 175).

| Sociodemographic Variables |  | *Sleeping Less Than Usual (N=884)* |  |  |  | *Sleeping More Than Usual (N=1903)* |  |  |  |
| --- | --- | --- | --- | --- | --- | --- | --- | --- | --- |
|  |  | *P-* |  |  |  | *P-* |  |  |  |
|  |  | *OR* | *Value* | *Lower* | *Upper* | *OR* | *Value* | *Lower* | *Upper* |
| Age | >70 | *Reference* |  |  |  |  |  |  |  |
|  | 60-69 | 1.33 | 0.117 | 0.93 | 1.89 | 1.23 | 0.103 | 0.96 | 1.57* |
|  | 50-59 | **2.05** | **0.000** | **1.39** | **3.02** | **1.48** | **0.006** | **1.12** | **1.96** |
|  | 40-49 | **2.24** | **0.000** | **1.45** | **3.46** | **1.58** | **0.005** | **1.15** | **2.17** |
|  | 30-39 | **2.92** | **0.000** | **1.86** | **4.58** | **1.62** | **0.005** | **1.15** | **2.27** |
|  | 18-29 | **2.10** | **0.013** | **1.17** | **3.78** | **2.18** | **0.000** | **1.42** | **3.36** |
| Sex | Male | *Reference* |  |  |  |  |  |  |  |
|  | Female | **1.86** | **0.000** | **1.56** | **2.22** | **1.20** | **0.007** | **1.05** | **1.37** |
| Race | White | *Reference* |  |  |  |  |  |  |  |
|  | Black, Hispanic, |  |  |  |  |  |  |  |  |
|  | Asian | 1.15 | 0.551 | 0.73 | 1.79 | 1.01 | 0.974 | 0.70 | 1.45 |
|  | Other | 0.72 | 0.154 | 0.45 | 1.13 | **0.55** | **0.002** | **0.37** | **0.80** |
| Child | No children | *Reference* |  |  |  |  |  |  |  |
|  | Child at home | 1.06 | 0.605 | 0.85 | 1.32 | 1.02 | 0.822 | 0.86 | 1.22 |
| Urban/Rural | Rural | *Reference* |  |  |  |  |  |  |  |
|  | Urban | **1.32** | **0.002** | **1.11** | **1.63** | **1.16** | **0.049** | **1.00** | **1.34** |
|  | Suburban | **1.35** | **0.027** | **1.03** | **1.77** | **1.24** | **0.038** | **1.01** | **1.52** |
| Marital Status | Married/cohabiting | *Reference* |  |  |  |  |  |  |  |
|  | Single | 1.09 | 0.468 | 0.86 | 1.40 | **1.27** | **0.012** | **1.05** | **1.53** |
|  | Widowed | 1.18 | 0.440 | 0.77 | 1.81 | 1.05 | 0.781 | 0.75 | 1.46 |
|  | Divorced/separated | **1.39** | **0.028** | **1.04** | **1.87** | **1.46** | **0.002** | **1.16** | **1.85** |
| Education | High School | *Reference* |  |  |  |  |  |  |  |
|  | Diploma or GED |  |  |  |  |  |  |  |  |
|  | College or Higher | 0.69 | 0.072 | 0.46 | 1.03 | **1.51** | **0.000** | **1.21** | **1.88** |
| Work | Employed | *Reference* |  |  |  |  |  |  |  |
|  | Unemployed | 1.21 | 0.250 | 0.87 | 1.68 | **1.80** | **0.000** | **1.41** | **2.30** |
|  | Student | 1.55 | 0.356 | 0.61 | 3.96 | **2.67** | **0.008** | **1.29** | **5.55** |
|  | Retired | 0.82 | 0.128 | 0.63 | 1.06 | **0.77** | **0.008** | **0.63** | **0.93** |
|  | Unable to work | 1.37 | 0.100 | 0.94 | 2.01 | 1.11 | 0.524 | 0.81 | 1.52 |
|  | Housework | 0.69 | 0.072 | 0.46 | 1.03 | **0.61** | **0.007** | **0.43** | **0.87** |
| Time Zone | Pacific | *Reference* |  |  |  |  |  |  |  |
|  | Mountain | 0.88 | 0.196 | 0.72 | 1.07 | **0.80** | **0.004** | **0.68** | **0.93** |
|  | Central | 0.89 | 0.481 | 0.65 | 1.23 | **0.72** | **0.012** | **0.56** | **0.93** |
|  | Eastern | 1.03 | 0.829 | 0.79 | 1.35 | 1.03 | 0.798 | 0.84 | 1.26 |

\* $\chi^2$ for trend=73.9, $p<0.0001$

Table 2 also shows associations of sociodemographic variables with sleeping more since hearing about the COVID-19 outbreak. There were 1,903 (36.7%) participants who slept more than usual. Age was an important associated factor, and analogous to sleeping less than usual, the odds of sleeping more appeared to become progressively greater with decreasing age. Similar to sleeping less, sleeping more was positively associated with female sex and being divorced or separated (vs. married). However, sleeping more also was related to being single in comparison to being married. Having a college education or higher was associated with more sleep compared to those with only a high school education. The impact of work status was mixed. In comparison to those who were employed, those who were unemployed or students slept more. In contrast, those who were retired or were homemakers were less likely to sleep more. The impact of time zone showed that those residing in the Mountain or Central time zones were less likely to sleep more. Although those whose race was other than Black, Hispanic or Asian appeared to be less likely to sleep more than Whites, the overall number of respondents to the survey in this demographic group was small.

Table 3 displays the association between COVID-19 beliefs and the likelihood of sleeping more or less since hearing about COVID-19. Sleeping less was associated with a higher perceived risk of COVID-19 infection, belief that COVID-19 was deadlier than the flu and belief that the problem was greater than what the government suggests. In contrast, more financial worries and disagreement with the statement that “COVID-19 is not as big a problem as the media suggest” was associated with lower odds of sleeping less. Sleeping more than usual was associated with the same pattern of responses as sleeping less than usual with one exception; there was a greater likelihood of sleeping more related to the belief that infection with COVID-19 would be associated with severe symptoms.

**Table 3.**
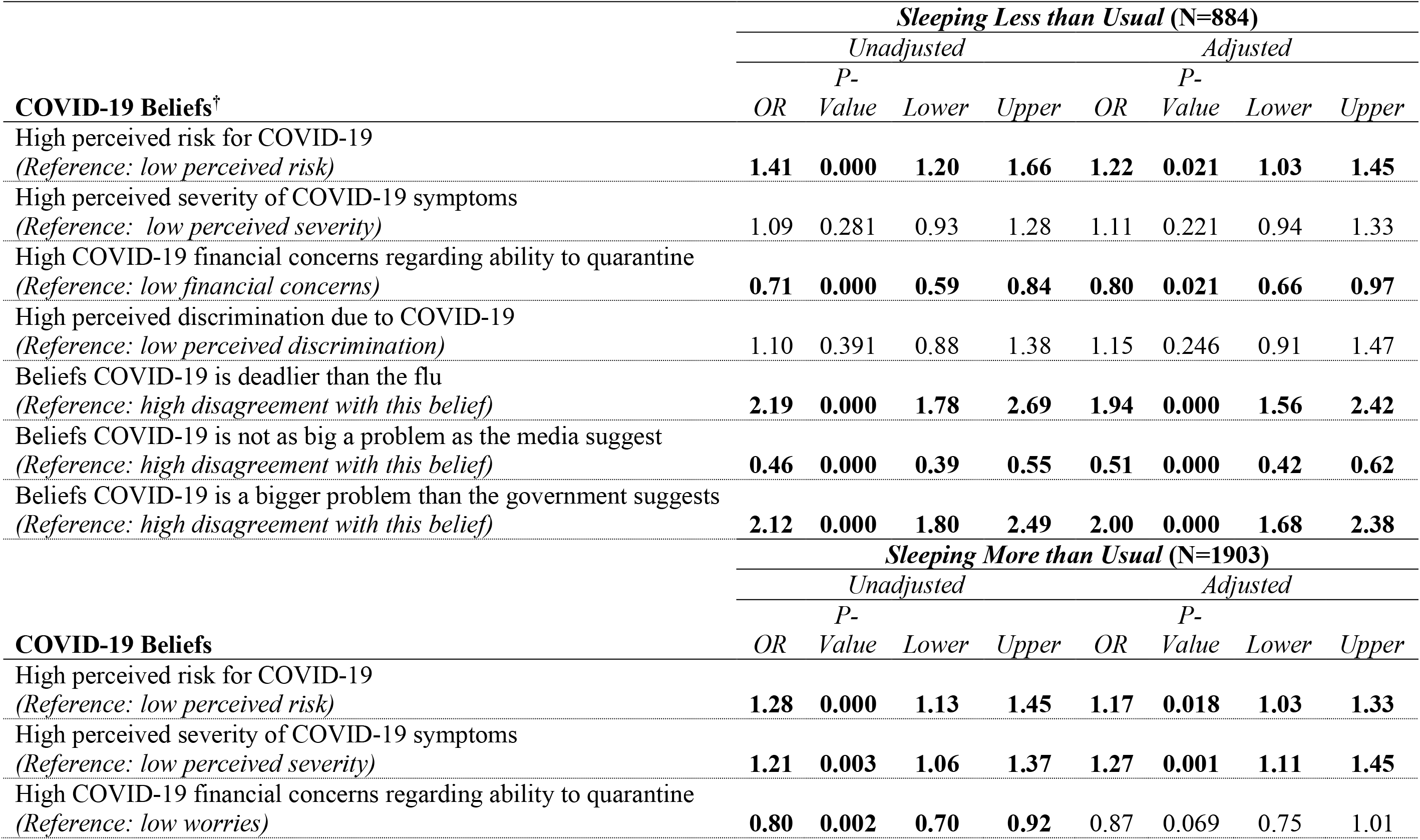
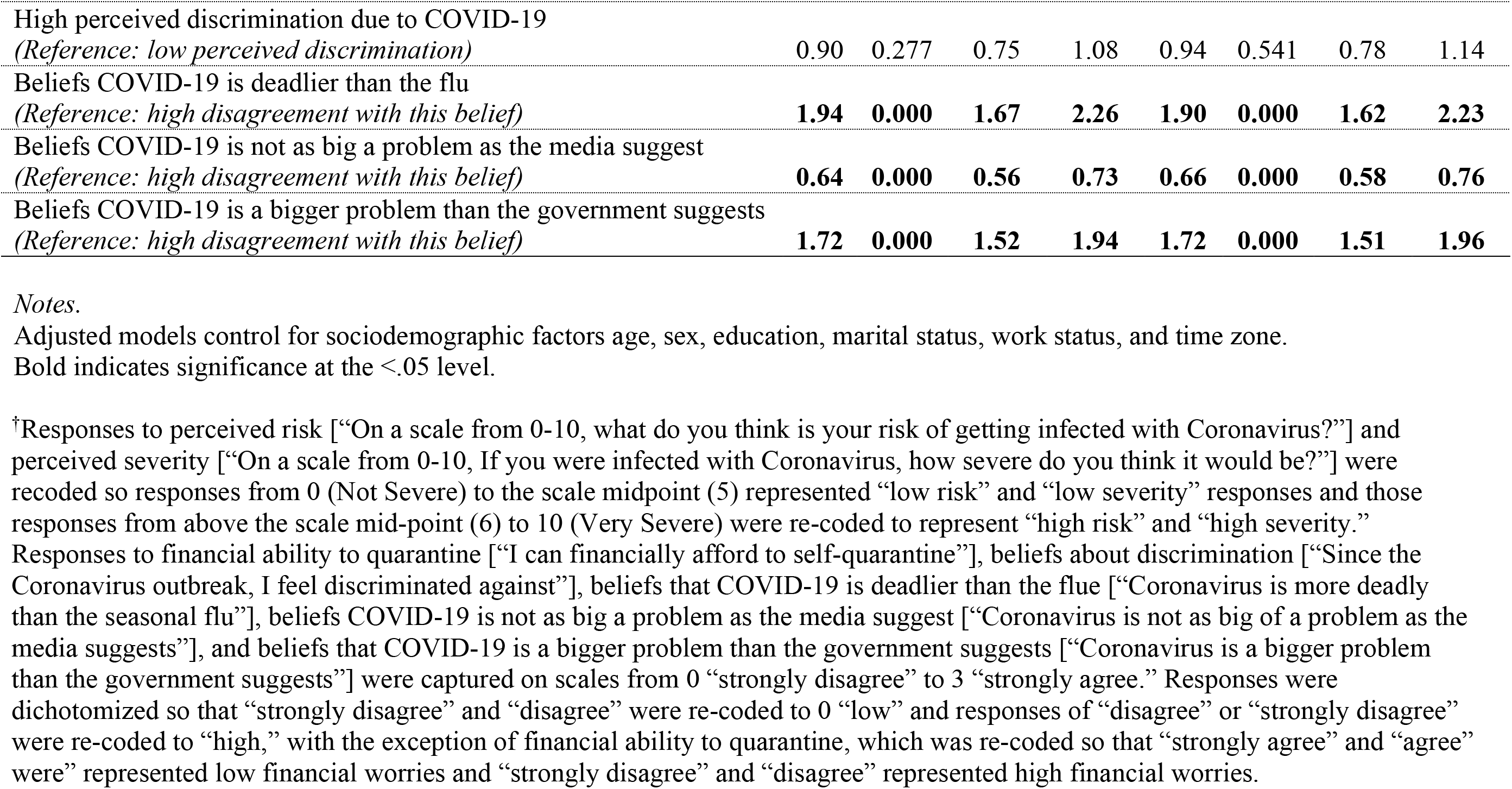
Binary Regression Examining the Relationships Between Sleeping Less or Sleeping More Than Usual and COVID-19 Beliefs in Adjusted and Unadjusted Models (N=5, 175).

| COVID-19 Beliefs<sup>†</sup> | *Sleeping Less than Usual (N=884)* |  |  |  |  |  |  |  |
| --- | --- | --- | --- | --- | --- | --- | --- | --- |
|  | *Unadjusted* |  |  |  | *Adjusted* |  |  |  |
|  | *OR* | *P-* | *Lower* | *Upper* | *OR* | *P-* | *Lower* | *Upper* |
|  |  | *Value* |  |  |  | *Value* |  |  |
| High perceived risk for COVID-19<br>(Reference: low perceived risk) | 1.41 | 0.000 | 1.20 | 1.66 | 1.22 | 0.021 | 1.03 | 1.45 |
| High perceived severity of COVID-19 symptoms<br>(Reference: low perceived severity) | 1.09 | 0.281 | 0.93 | 1.28 | 1.11 | 0.221 | 0.94 | 1.33 |
| High COVID-19 financial concerns regarding ability to quarantine<br>(Reference: low financial concerns) | 0.71 | 0.000 | 0.59 | 0.84 | 0.80 | 0.021 | 0.66 | 0.97 |
| High perceived discrimination due to COVID-19<br>(Reference: low perceived discrimination) | 1.10 | 0.391 | 0.88 | 1.38 | 1.15 | 0.246 | 0.91 | 1.47 |
| Beliefs COVID-19 is deadlier than the flu<br>(Reference: high disagreement with this belief) | 2.19 | 0.000 | 1.78 | 2.69 | 1.94 | 0.000 | 1.56 | 2.42 |
| Beliefs COVID-19 is not as big a problem as the media suggest<br>(Reference: high disagreement with this belief) | 0.46 | 0.000 | 0.39 | 0.55 | 0.51 | 0.000 | 0.42 | 0.62 |
| Beliefs COVID-19 is a bigger problem than the government suggests<br>(Reference: high disagreement with this belief) | 2.12 | 0.000 | 1.80 | 2.49 | 2.00 | 0.000 | 1.68 | 2.38 |
| COVID-19 Beliefs | *Sleeping More than Usual (N=1903)* |  |  |  |  |  |  |  |
|  | *Unadjusted* |  |  |  | *Adjusted* |  |  |  |
|  | *OR* | *P-* | *Lower* | *Upper* | *OR* | *P-* | *Lower* | *Upper* |
|  |  | *Value* |  |  |  | *Value* |  |  |
| High perceived risk for COVID-19<br>(Reference: low perceived risk) | 1.28 | 0.000 | 1.13 | 1.45 | 1.17 | 0.018 | 1.03 | 1.33 |
| High perceived severity of COVID-19 symptoms<br>(Reference: low perceived severity) | 1.21 | 0.003 | 1.06 | 1.37 | 1.27 | 0.001 | 1.11 | 1.45 |
| High COVID-19 financial concerns regarding ability to quarantine<br>(Reference: low worries) | 0.80 | 0.002 | 0.70 | 0.92 | 0.87 | 0.069 | 0.75 | 1.01 |
| High perceived discrimination due to COVID-19<br>(Reference: low perceived discrimination) | 0.90 | 0.277 | 0.75 | 1.08 | 0.94 | 0.541 | 0.78 | 1.14 |
| Beliefs COVID-19 is deadlier than the flu<br>(Reference: high disagreement with this belief) | **1.94** | **0.000** | **1.67** | **2.26** | **1.90** | **0.000** | **1.62** | **2.23** |
| Beliefs COVID-19 is not as big a problem as the media suggest<br>(Reference: high disagreement with this belief) | **0.64** | **0.000** | **0.56** | **0.73** | **0.66** | **0.000** | **0.58** | **0.76** |
| Beliefs COVID-19 is a bigger problem than the government suggests<br>(Reference: high disagreement with this belief) | **1.72** | **0.000** | **1.52** | **1.94** | **1.72** | **0.000** | **1.51** | **1.96** |

Table 4 shows the relationship between mental health constructs and changes in sleep duration. Both sleeping less than usual and sleeping more than usual were strongly associated with anxiety, depression, and loneliness in unadjusted and fully adjusted models.

**Table 4.**
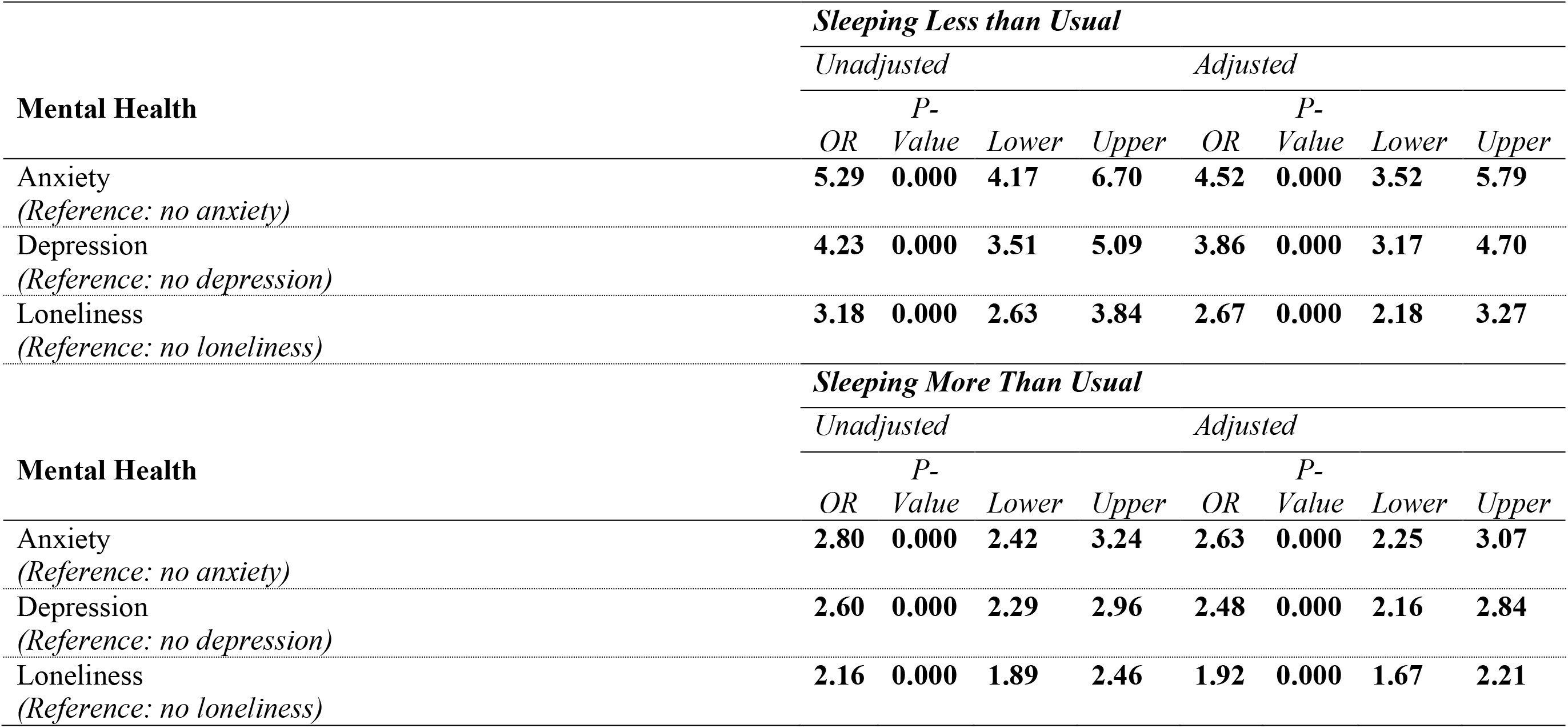
Binary Regression Variables Examining the Relationships Between Sleeping Less or Sleeping More Than Usual and Mental Health in Adjusted and Unadjusted Models.

|  |  | ***Sleeping Less than Usual*** |  |  |  |  |  |  |  |
| --- | --- | --- | --- | --- | --- | --- | --- | --- | --- |
|  |  | *Unadjusted* |  |  |  | *Adjusted* |  |  |  |
| **Mental Health** |  | *OR* | *P-Value* | *Lower* | *Upper* | *OR* | *P-Value* | *Lower* | *Upper* |
| Anxiety |  | **5.29** | **0.000** | **4.17** | **6.70** | **4.52** | **0.000** | **3.52** | **5.79** |
| *(Reference: no anxiety)* |  |  |  |  |  |  |  |  |  |
| Depression |  | **4.23** | **0.000** | **3.51** | **5.09** | **3.86** | **0.000** | **3.17** | **4.70** |
| *(Reference: no depression)* |  |  |  |  |  |  |  |  |  |
| Loneliness |  | **3.18** | **0.000** | **2.63** | **3.84** | **2.67** | **0.000** | **2.18** | **3.27** |
| *(Reference: no loneliness)* |  |  |  |  |  |  |  |  |  |
|  |  | ***Sleeping More Than Usual*** |  |  |  |  |  |  |  |
|  |  | *Unadjusted* |  |  |  | *Adjusted* |  |  |  |
| **Mental Health** |  | *OR* | *P-Value* | *Lower* | *Upper* | *OR* | *P-Value* | *Lower* | *Upper* |
| Anxiety |  | **2.80** | **0.000** | **2.42** | **3.24** | **2.63** | **0.000** | **2.25** | **3.07** |
| *(Reference: no anxiety)* |  |  |  |  |  |  |  |  |  |
| Depression |  | **2.60** | **0.000** | **2.29** | **2.96** | **2.48** | **0.000** | **2.16** | **2.84** |
| *(Reference: no depression)* |  |  |  |  |  |  |  |  |  |
| Loneliness |  | **2.16** | **0.000** | **1.89** | **2.46** | **1.92** | **0.000** | **1.67** | **2.21** |
| *(Reference: no loneliness)* |  |  |  |  |  |  |  |  |  |

## Discussion

This study examined changes in sleep duration among a national sample of social media users since hearing about the COVID-19 pandemic as well as factors associated with these changes. We demonstrated that a large proportion of the United States adult population suffered a change in sleeping patterns during this pandemic. Importantly, these changes disproportionately impacted women, younger to middle aged adults, and those living in urban settings. Differential associations were further noted as a consequence of marital status, employment, and education. Being single and employed was associated with sleeping less than usual. However, the strongest associations were observed with symptoms of mental health impairment.

One of the most important findings in our analyses was that 53.9% of our cohort experienced a change in their sleep duration as a result of the COVID-19 pandemic with the largest proportion (36.7%) sleeping more than usual. This latter finding is consistent with recent data from a global internet survey which demonstrated an average 26 minute increase in sleep duration and 50 minute delay in mid-sleep time on workdays^28^ and from another survey using data downloaded from a widely used sleep app showing a mean increase of 14 minutes in 5 large American, European and Asian cities,^29^ as well as an internet survey in Americans.^30^ The ability to work from home without “social time pressure” is the most likely explanation.^28^ Moreover, given the multitude of stressors during this pandemic, fatigue and depression may also be contributing to increased sleep time.

Notwithstanding the large percentage of those who increased their sleep duration, there also were 17.1% of participants who were sleeping less than usual. This finding of worsening sleep during the pandemic is consistent with previous studies demonstrating increased rates of insomnia. In mainland China, an online survey of participants between the ages of 18-76 years found that the prevalence of insomnia rose from 26.2% before to 33.7% during the pandemic.^31^ Similarly, a recent meta-analysis also demonstrated a high prevalence of insomnia (23.8%) with a greater impact among healthcare workers in comparison to the general population.^32^ In another meta-analysis,^33^ researchers reported an overall pooled prevalence of insomnia of over 37% with higher rates among healthcare workers compared to the general population (47.3% vs 18.2%). Other studies have reported poor sleep among healthcare workers during previous SARS and MERS pandemics.^34,35^ However, most of these studies are from China and Europe where the devastation from this pandemic was felt initially and without prior knowledge of the overall impact of this virus. The uncertainty surrounding the pandemic likely contributed to stress,^36^ a major contributor to insomnia.

Our study found that women were more likely to report both sleeping less and sleeping more in response to the current pandemic. Our results are consistent with those reported in a Greek population.^37^ Women maybe more vulnerable to stress, and in general report more somatic symptoms and poor sleep than men.^33^ As evidenced in our study, sleep duration is also impacted more by childcare needs which has a greater consequence for women. Additionally, most studies demonstrating higher prevalence of insomnia among healthcare workers have a higher proportion of women in their cohorts; insomnia generally is more prevalent in women.^38^

The other major demographic factor associated with a change in sleep duration was age. For both sleeping more and for sleeping less, we found that the impact of the pandemic was greater in younger persons. To our knowledge this is the first study demonstrating increased sleep among older participants during this crisis. This observation is in contradiction to the known association of age with the greater likelihood of having a sleep disturbance.^39^ It is possible that financial security among the elderly lead to less worries and anxiety and hence less risk of sleep disturbance despite greater morbidity and mortality risk from COVID-19. Furthermore, in a previous influenza epidemic, high stress levels were observed disproportionately in younger adults.^40^ It is possible that younger persons are more vulnerable and have lesser coping skills than older participants. Consistent with this explanation, Huang et al. have reported elevated levels of anxiety among young adults during the pandemic.^41^

We found that those living in urban and suburban settings were more likely to experience a change in sleep duration. Our findings are consistent with a study demonstrating greater sleep disturbances among urban Greek residents during the pandemic compared to those living in rural settings.^37^ The fact that the first phase of the COVID-19 pandemic primarily affected the more densely populated urban and suburban areas of the United States is the most likely explanation for this finding in our study. It also is consistent with previous observations demonstrating that sleep quality generally is worse among urban residents.^42^ Rural living also may be protective by providing more opportunities for safe physical activity and exposure to natural environments resulting in less stress.^37^

Several other sociodemographic factors appeared to play a role in altering sleep duration. Some categories of living alone were a risk factor for less sleep or more sleep suggesting that the presence of a partner mitigated some of the pandemic-related social isolation. Education was associated with sleeping more. It is likely that higher levels of education provided more opportunity to work from home.^43^ Being unemployed or a student increased the likelihood of sleeping more; absence of “social time pressure” is a likely explanation. Being a homemaker or retired was not associated with sleeping more. The former may reflect the realities of the high workload of a homemaker. The latter may be either financially secure or are less likely to be affected by changes in income / loss of income and employment due to COVID-19. An interesting finding was the impact of time zone with participants residing in the Mountain or Central time zones less likely to sleep more. A previous study has demonstrated that Mountain or Central time zone residents sleep on average 15 minutes more per night possibly related to scheduling of television network programming (1 hour earlier in these time zones).^44^ A pre-existing longer sleep duration may have made these participants less vulnerable to increasing their sleep duration further.

We observed that both sleeping more and sleeping less were highly associated with beliefs related to the impact of COVID-19 on personal health or financial status. Individuals who felt that COVID-19 would negatively impact them were more likely to have a change in sleep duration. Our results are consistent with those reported from the United Kingdom with respect to increased risk of being infected with COVID-19.^9^ The likely explanation for these observations is greater stress levels and their negative impact on sleep. However, the finding that greater worry regarding the ability to quarantine was associated with a lower association with less sleep seems counterintuitive and without an obvious explanation. Interestingly, despite reports of adverse racially motivated actions perpetrated against Asians, there was no impact of high levels of perceived discrimination on sleep duration. However, the number of Asians in the study cohort was very low, precluding us from drawing conclusions with regards to race and ethnicity.

Mental health constructs appeared to be the most highly associated factors with both more sleep and less sleep. The association between depression and anxiety, and insomnia during the COVID-19 pandemic is well-established in both health care workers and the general population.^16,33^ We extend these findings by demonstrating that both depression and anxiety are related to both more sleep and less sleep. Surveys during the pandemic have also documented that loneliness occurs commonly and is related to the social isolation imposed by COVID-19 mitigation strategies.^45,46^ Our findings documenting the association between loneliness and changes in sleep duration highlight the impact of social isolation on sleep. It is possible that interventions to improve mental health during the pandemic will result in better sleep health.

A major strength of this study is that it utilized a dataset containing substantial information pertaining to sociodemographic and mental health status that might explain change in sleep duration. However, we do acknowledge some limitations. First, the study recruitment was comprised mainly of a convenience sample of online social media users. Although 70% of Americans use Facebook,^47^ certain demographic groups may have been underrepresented. Nevertheless, a recent systematic review has identified Facebook as a useful tool in shortening recruitment periods, reducing costs, and enhancing representation of target populations.^20^ Second, people with low socioeconomic status and poor access to computer or internet connection (11.2%-19.6%)^48^ at home also may have been underrepresented in this study. Although we tried to recruit racial and ethnic minorities,^18^ we do acknowledge that our sample was predominantly white and not representative of the United States population. However, a mitigating factor is that we had respondents from every state, age group and sex. Unfortunately, given social distancing regulations caused by the COVID-19 pandemic, online data collection is one of the few platforms that is available. Third, our data regarding sleep was limited only to a single self-reported question on changes in sleep duration. It is therefore relatively non-specific and carries the risk of recall bias. In addition, we lack information on COVID-19 infection status for study participants; this maybe important information since sleep disturbances can be a consequence of viral infection. Lastly, our analyses were cross-sectional, and thus causality cannot be inferred.

In summary, we have presented data showing that during the COVID-19 pandemic, the majority of adults in the United States experience either an increase or a decrease in their sleep and that changes in sleep duration are related to a number of sociodemographic factors and beliefs regarding susceptibility to COVID-19 infection. Importantly, mental health constructs have the strongest association with sleep suggesting that in addition to prevention and treatment of COVID-19, recognizing psychosocial aspects during stressful times is crucial. Future studies are needed to examine this association between COVID-19 and neuropsychiatric problems, and to develop interventions to address these challenges across populations resulting in better sleep health.

## Data Availability

Requests for data used in this study should be made to the corresponding author.

## Acknowledgments

The authors respectfully acknowledge the contribution of Ms. Kristen Monten at an earlier stage of this manuscript. This work was supported in part by the NIH grant numbers: K01HL150339 (RR)

## Supplement

1. Sex
  a. Female
  b. Male
  c. Other
  d. Prefer not to disclose
2. Age group
  a. 18-29 years old
  b. 30-39 years old
  c. 40-49 years old
  d. 50-59 years old
  e. 60-69 years old
  f. 70-79 years old
  g. 80+ years old
3. What’s your race/ethnicity?
  a. Black, Non-Hispanic
  b. White, Non-Hispanic
  c. Asian/Pacific Islander
  d. Hispanic/Latinx
  e. Native American or American Indian
  f. Interracial, Mixed race, or Other
4. What is your marital or cohabitation status?
  a. Married/cohabitating
  b. Single
  c. Widowed
  d. Divorced/separated
5. How would you describe where you live?
  a. Urban
  b. Suburban
  c. Rural
6. What is your current working status?
  a. Employed, full time
  b. Employed, part time
  c. Unpaid work (e.g., homemaker, eldercare, childcare)
  d. Self-employed
  e. Out of work and looking for work
  f. Out of work but not currently looking for work
  g. Student
  h. Military personnel
  i. Retired
  j. Unable to work
7. What is your highest level of education?
  a. Less than a High School diploma
  b. High School diploma or GED
  c. Some college, no degree
  d. Associate’s degree
  e. Bachelor’s degree
  f. Masters/Professional degree or above
8. In which state do you currently reside? [Drop-down menu of all 50 states, District of Columbia, Puerto Rico]
9. What do you think is your risk of getting infected with the Coronavirus?

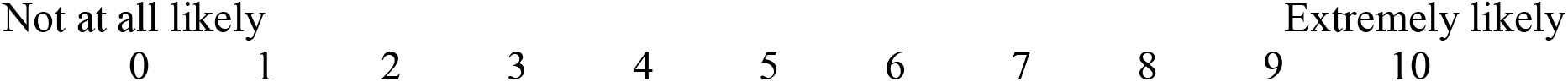
10. If you were infected with the Coronavirus, how severe do you think it would be?

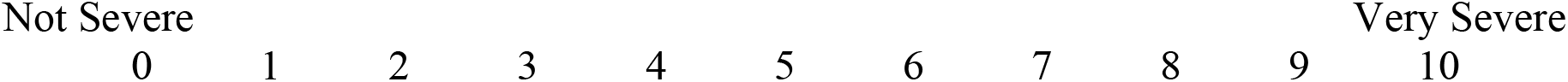
11. I can financially afford to self-quarantine.
  a. Strongly Agree
  b. Agree
  c. Disagree
  d. Strongly Disagree
12. Since the Coronavirus outbreak, I feel discriminated against.
  a. Strongly Agree
  b. Agree
  c. Disagree
  d. Strongly Disagree
13. Coronavirus is more deadly than the seasonal flu.
  a. Strongly Agree
  b. Agree
  c. Disagree
  d. Strongly Disagree
14. Coronavirus is not as big of a problem as the media suggests.
  a. Strongly Agree
  b. Agree
  c. Disagree
  d. Strongly Disagree
15. Coronavirus is a bigger problem than the government suggests.
  a. Strongly Agree
  b. Agree
  c. Disagree
  d. Strongly Disagree
  *In the last 3 months, how often have you felt:*
16. Left out
  a. Never
  b. Rarely
  c. Sometimes
  d. Often
17. Isolated from others
  a. Never
  b. Rarely
  c. Sometimes
  d. Often
18. That you lack companionship
  a. Never
  b. Rarely
  c. Sometimes
  d. Often
  *Over the last 7 days, how often have you been bothered by any of the following problems because of THE CORONAVIRUS OUTBREAK?*
19. Feeling nervous, anxious, or on edge?
  a. Not at all
  b. Several days
  c. More than half the days
  d. Nearly everyday
20. Not being able to stop or control worrying?
  a. Not at all
  b. Several days
  c. More than half the days
  d. Nearly everyday
21. Feeling down, depressed, or hopeless?
  a. Not at all
  b. Several days
  c. More than half the days
  d. Nearly everyday
22. Little interest or pleasure in doing things (that I used to enjoy)?
  a. Not at all
  b. Several days
  c. More than half the days
  d. Nearly everyday
  *Since hearing about the Coronavirus outbreak, how have the below behaviors changed for you?*
23. Sleeping
  a. Much more
  b. Little more
  c. Not changed
  d. Little less
  e. Much less
  f. Not applicable

